# Identification of rare missense variants reducing cathepsin O secretion in families with intracranial aneurysm

**DOI:** 10.1101/2023.01.31.23285168

**Authors:** Milène Freneau, Raphael Blanchet, Sandro Benichi, Mary-Adel Mrad, Surya Prakash Rao Batta, Marc Rio, Stéphanie Bonnaud, Pierre Lindenbaum, Fabien Laporte, Stéphane Cuénot, Thibaud Quillard, Sandrine Morel, Brenda R. Kwak, Philippe Bijlenga, Jean-François Deleuze, Christian Dina, Maxence Bodet, Stéphanie Chatel, Emmanuelle Bourcereau, Solène Jouan, Arturo Consoli, Cyril Dargazanli, Julien Ognard, ICAN Study Group, Hubert Desal, Anne-Clémence Vion, Romain Bourcier, Gervaise Loirand, Richard Redon

## Abstract

Intracranial aneurysm (IA) is a common cerebrovascular abnormality characterized by localized dilation and wall thinning in intracranial arteries, which can rupture and lead to fatal subarachnoid hemorrhage. Although the pathophysiology of IA remains largely unknown, increasing evidence suggests that genetic susceptibility plays a predominant role. Here, we combined whole exome sequencing and identity-by-descent analyses with functional investigations to identify rare functional variants associated to IA in families with multiple affected subjects. We identified two rare missense variants in the *CTSO* gene in two large pedigrees. We found that the cysteine-type papain-like cathepsin O (CTSO) encoded by *CTSO* is expressed in the wall of human IA domes. Stretching of vascular smooth muscle cells (VSMC) induced secretion of CTSO, which acted as an extracellular protease controlling VSMC migration and adhesion to the extracellular matrix. CTSO depletion, as well as expression of the two CTSO variants, which are poorly secreted, increased the amount of fibronectin. In addition, CTSO depletion increased VSMC stiffness, which was reduced by the addition of exogenous CTSO. Collectively, our findings identify CTSO as a potential new player in arterial remodeling, regulating fibronectin deposition and VSMC function, supporting the causal role of rare coding *CTSO* variants in familial forms of IA.

## INTROCUCTION

Intracranial aneurysm (IA) is a generally asymptomatic localized dilation associated with thinning and structural defects of the intracranial arterial wall affecting 1-5% of the general population (1). The main risk of IA is its unpredictable rupture, responsible for ∼36,000 of subarachnoid hemorrhage cases per year in Europe and a major cause of sudden death in “young healthy” subjects, in particular in women (peak of age: 50-60 years) (2, 3). Although hypertension, ageing, female sex, smoking, excessive alcohol consumption and family history of IA have been identified as risk factors predisposing to IA, the mechanisms underlying IA formation, growth and rupture are mostly unknown. However, local hemodynamic constraints of shear stress and pressure are assumed to play a key role (4). The aggregation of patients in IA families suggests that genetic factors contribute to disease susceptibility, and identifying these factors might be beneficial for at-risk patients before IA rupture (5, 6). Genome-wide association studies have identified several common risk loci associated with IA (7–10). However, the identification of specific genes or causal molecular pathways remains largely inconclusive and these common alleles together only explain a small fraction of the risk attributable to genetics for IA in the general population. On the other hand, whole-exome sequencing (WES) has the potential to detect rare coding variants that have large effect and induce a high risk of developing IA. Identifying such variants also could provide valuable insights into IA pathophysiology and pave the way for new diagnostic or therapeutic strategies. Indeed, recent studies have demonstrated the usefulness of familial approaches based on WES to improve knowledge on the molecular mechanisms underlying IA formation and rupture from the discovery of rare predisposing variants such as *RNF213* (11), *THSD1* (12), *LOXL2* (13), *PCNT* (14), *ARHGEF17* (15), *ANGPTL6* (16), and *PPIL4* (17).

In the present study, by combining WES and identity-by-descent (IBD) analysis, we identified rare coding variants in the Cathepsin O gene (*CTSO* [MIM: 600550]) in two familial cases of IA. CTSO is one of the 11 cysteine-type papain-like cathepsins identified in humans about which almost nothing is known (18). Our functional analysis revealed that CTSO controls vascular smooth muscle cell (VSMC) migration and adhesion to the extracellular matrix (ECM). Furthermore, CTSO depletion increased VSMC stiffness. These finding suggest that the increased susceptibility to IA induced by *CTSO* variants is likely related to their primary effects on vascular tissue, and more particularly on the media layer of the wall of cerebral arteries.

## RESULTS

### Identification of one rare coding variant affecting *CTSO* in a familial case of IA

We first recruited a large family (pedigrees are available on request) with 6 definite carriers of IA in two generations, among whom two had a history of IA rupture. In this family, there were also 4 relatives with suspected IA (3 because of unexplained sudden death and 1 with a doubtful ectasia on MRI, see Figure S1A), 3 individuals with unknown phenotypes in the absence of MRI screening, and 19 subjects without IA detected on MRI. Among the definite carriers of IA, all were females and 3 were carriers of multiple IA (with a maximum of 4 IAs detected for 1 subject). Of the twelve IAs detected in the family, 6 were located in the MCA territory (50%), 3 in the ACA (25%), 2 in the PCA (16%), and 1 in the ICA territory (9%)(Figure 1A and B). The median maximal diameter of IA was 4 mm (min 3 mm, max 8 mm). We applied whole-exome sequencing to 3 affected members of the family. Out of the 77 to 107 rare non-synonymous or splicing variants detected in each 3 whole exomes, we found that only 8 were shared between the 3 affected relatives (Table S1 and S2). IBD analysis on all available relatives revealed that only one of the 8 variants resides within a shared haplotype - located at 4q32 - between the 6 affected subjects (Table S1 and S3). We then confirmed this finding by capillary sequencing. This missense variant in the *CTSO* gene - *CTSO* c.946G>A (p.Val316Ile) – is reported with a Genomic Evolutionary Rate Profiling (GERP) score of 5.68 and predicted *in silico* as damaging for CTSO protein structure and function by both PolyPhen-2 and SIFT (Table 1).

**Figure 1.**
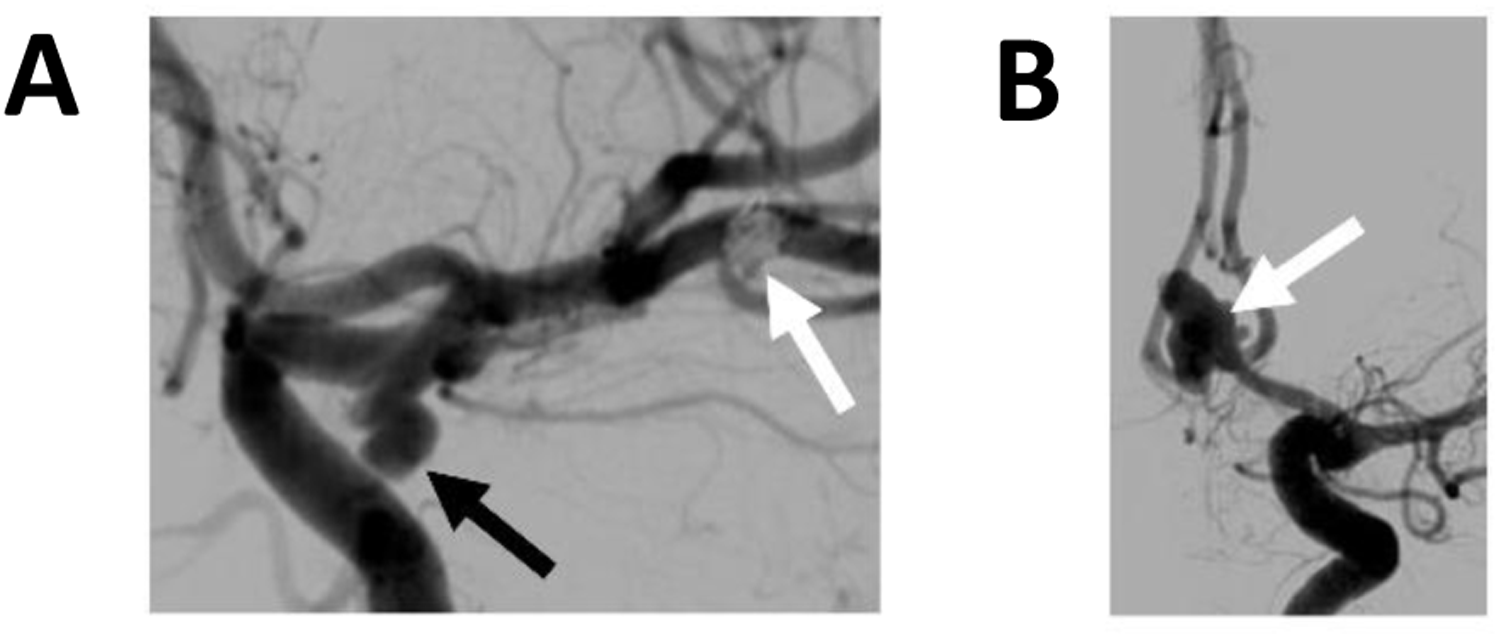
Angiography of two subjects from families with IA. (**A**) Digital subtracted angiography showing two IA carried by the index case II-9 of family A: one located on the middle cerebral artery and already treated by coiling (white arrow), and one located on the posterior communicating artery (black arrow). (**B**) Digital subtracted angiography showing one ruptured ICA just before treatment, located on the anterior communicating artery (white arrow).

**Table 1.** Rare functional variants in *CTSO* shared by all IA-affected subjects in families A and B.

| Family | Genomic position<br>(GRCh37/hg19) | Gene | Nucleotide<br>consequence | Protein<br>consequence | MAF (NFE in<br>gnomAD) | Predicted Functional Impact |  | Cases / Total |  |
| --- | --- | --- | --- | --- | --- | --- | --- | --- | --- |
|  |  |  |  |  |  | SIFT | Polyphen 2 | Affected | Unaffected |
| A | 4:156847208 | *CTSO* | c.946G>A | p.Val316Ile | $1.7 \cdot 10^{-5}$ | Deleterious | Probably<br>damaging | 6/6 | 8/19 |
| B | 4:156874872 | *CTSO* | c.128C>T | p.Ala43Val | $3.0 \cdot 10^{-4}$ | Tolerated | Probably<br>damaging | 3/3 | 0/5 |

Mutation names are based on Ensembl *CTSO* transcript ENST00000433477.3. MAF: Minor
Allele Frequency; NFE: Exome Aggregation Consortium (Non-Finnish Europeans);
GnomAD: genome Aggregation Database; GERP: Genomic Evolutionary Rate Profiling;
SIFT: Sorting Intolerant From Tolerant.

### A second familial case of IA associated with rare *CTSO* coding variant

To further investigate the putative involvement of *CTSO* in IA susceptibility, we then screened for rare functional variants within this gene among 93 additional index cases with familial forms of IA, for which WES data were available in-house. We found one additional rare missense variant, *CTSO* c.128C>T (p.Ala43Val), in one case (Table 1). The corresponding family comprised 4 IA carriers (3 females and 1 males), among whom one carried 2 IAs. The median maximal diameter of IA was 5 mm (min 4 mm, max 9 mm). Three IAs were located in the MCA territory, one in the ACA and one in the ICA territory, while in one case the location was undetermined. undetermined. By capillary sequencing, we tested the *CTSO* c.128C>T variant among family members and detected it in the 3 tested IA carriers but in none of their 6 non-affected relatives (including a subject with uncertain phenotype, Figure S1B). This variant, reported with a GERP score of 3.67, was predicted *in silico* as damaging by PolyPhen-2 but not by SIFT (Table 1).

### Clinical characteristics and exposition to risk factors according to *CTSO* status

When comparing subgroups of individuals with or without *CTSO* variant and carrying or not IA among the two families members, we observed a higher proportion of subjects with history of high blood pressure among the IA carriers (p=0.013) while we found no significant differences regarding age, proportion of women, smoking habits, history of ischaemic stroke, level of alcohol consumption, history of diabetes or dyslipidaemia, treatment with statin, antiplatelet, oral anticoagulant and anti-inflammatory medication (Table 2).

**Table 2.** Clinical characteristics, treatments and exposition to risk factors for IA cases and unaffected relatives according to *CTSO* status.

|  | ***CTSO* variants**<br>**IA** | ***CTSO* variants**<br>**no IA** | **No *CTSO* variant**<br>**no IA** | ***P*-value** |
| --- | --- | --- | --- | --- |
| n | 9 | 8 | 16 | - |
| Age (yr) [median (range)] | 69 (51 - 93) | 60 (38 - 89) | 49 (33 - 76) | - |
| Female | 8 (88.9%) | 3 (37.5%) | 9 (56.3%) | 0.088 |
| Smoker or former smoker | 4 (44.4%) | 5 (62.5%) | 8 (50%) | 0.816 |
| Alcohol intake<br>> 150 g per week | 4 (44.4%) | 1 (12.5%) | 3 (18.8%) | 0.326 |
| High Blood Pressure | 5 (55.6%) | 2 (25%) | 1 (6.3%) | 0.013 |
| Dyslipemia or diabetes | 2 (22.2%) | 2 (25%) | 0 (0%) | 0.080 |
| Antiplatelet treatment | 2 (22.2%) | 1 (12.5%) | 1 (6.3%) | 0.666 |
| Anti-inflammatory drugs | 0 (0%) | 0 (0%) | 1 (6.3%) | 1.000 |
| Statin treatment | 1 (11.1%) | 1 (12.5%) | 1 (6.3%) | 1.000 |
| Oral anticoagulant<br>treatment | 1 (11.1%) | 2 (25%) | 4 (25%) | 0.745 |

Data are expressed as median, (minimum, and maximum) for age and as count (and percentage) for all other categorical variables. The Fischer exact probability test was used for contingency comparison between groups.

### CTSO is expressed in cerebral arteries

To explore the putative implication of CTSO in IA pathophysiology, we first sought to investigate the expression of CTSO in cerebral arteries. *Ctso* mRNA was found at a similar level in cerebral arteries of WKY rats and in arteries from spontaneously hypertensive rat (SHR) and stroke prone SHR (SPSHR), suggesting that *Ctso* mRNA expression was not modulated by high blood pressure (Figure 2A). In cultured cells, *Ctso* mRNA was expressed in VSMC (Figure 2B) and, at weaker levels, in endothelial cells, where it was not affected by shear stress (SS) (Figure 2C, D). In VSMC, CTSO was found in cell lysates but also in the culture medium, indicating the secretion of the protein (Figure 2E). Moreover, CTSO secretion was potentiated by stretch, without any change in mRNA level (Figure 2B and Figure 2E). Interestingly, CTSO is expressed in the arterial wall of human IA domes (Figure 3A). In unruptured IA, CTSO was expressed in some endothelial cells and VSMC, but CTSO staining was not observed in ECM. A significantly higher content of CTSO, particularly in the ECM was detected in the arterial wall of ruptured IA thus confirming CTSO secretion in the extracellular space by VSMC observed *in vitro*, particularly under stretching condition (Figure 3B).

**Figure 2.**
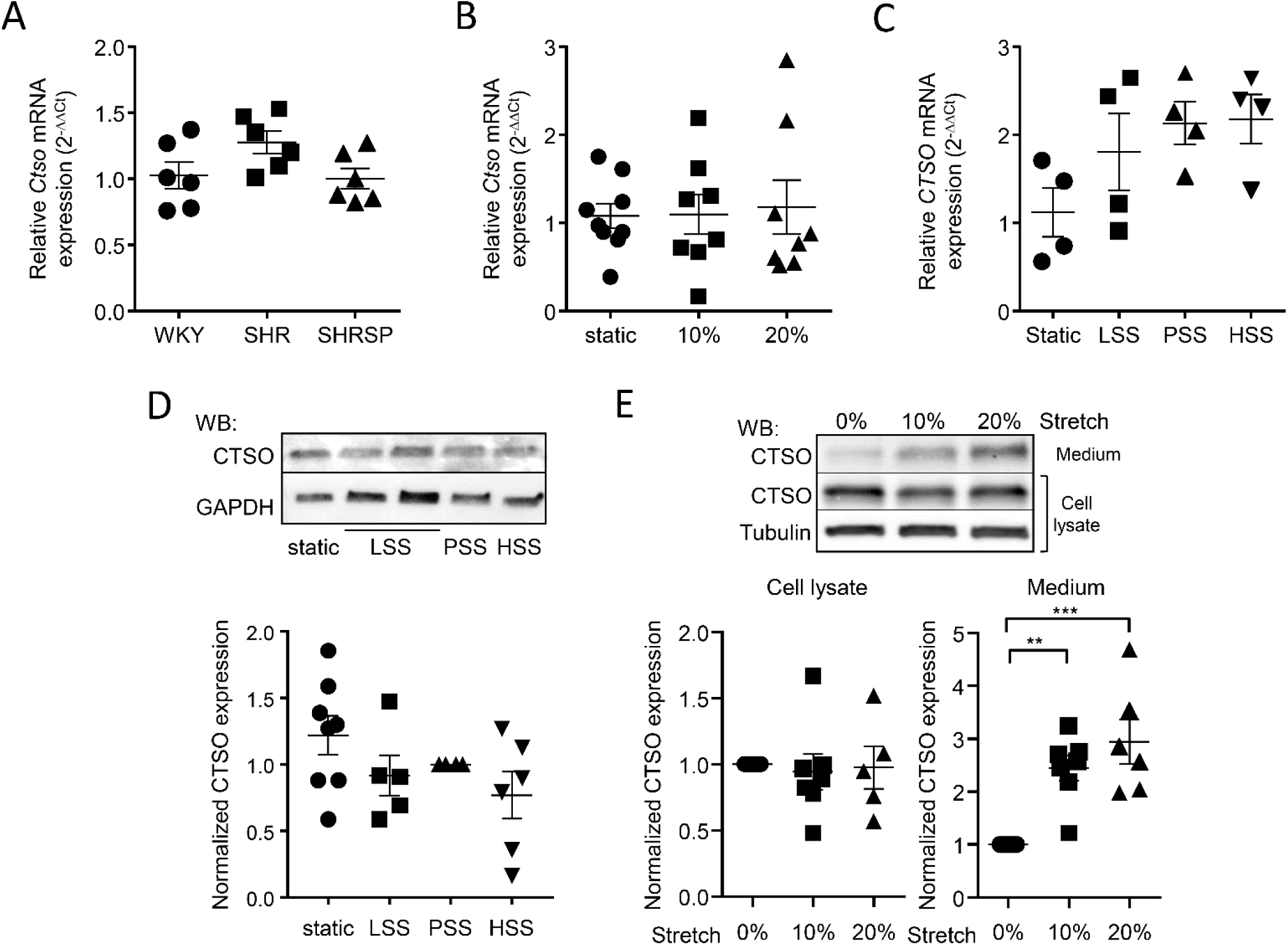
CTSO expression in arteries, smooth muscle and endothelial cells. (**A**) *Ctso* mRNA expression analyzed by quantitative RT-PCR in cerebral arteries of normotensive (WKY), and hypertensive rats (SHR and SHRSP). (**B**) *Ctso* mRNA expression analyzed by quantitative RT-PCR in VSMC cultured in static condition and subjected to 10% and 20% cyclic stretch. (**C**) *CTSO* mRNA expression analyzed by quantitative RT-PCR in HUVEC under static condition and under flow generating low (LSS, 3 dyn/cm^2^), physiological (PSS, 16 dyn/cm^2^) and very high (HSS, 36 dyn/cm^2^) shear stress. Means ± SEM are shown. (**D**) CTSO expression in HUVEC under static condition and under flow (LSS, PSS and HSS) and representative Western blots. GAPDH was also blotted to check equal loading. (**E**) Representative western blots and quantification of CTSO protein expression assessed in cell lysate and culture medium of VSMC cultured in static condition and subjected to 10% and 20% cyclic stretch. (Data are expressed as mean ± SEM; \*\**p*<0.01 and \*\*\**p*<0.001; one-way *ANOVA*).

**Figure 3.**
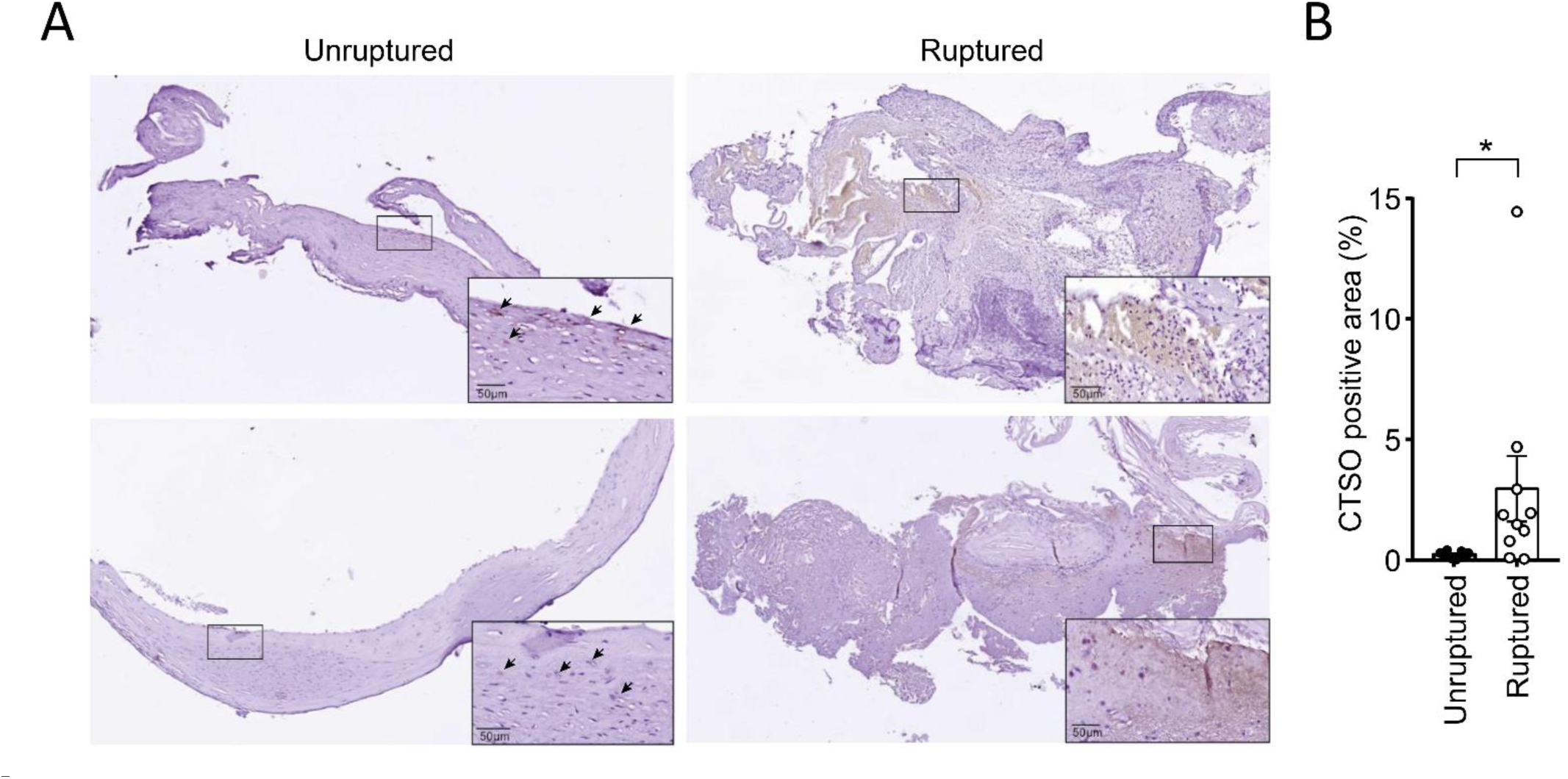
Expression of CTSO in human IA domes. (**A** and **B**) Representative whole images of immunochemical staining of CTSO (in brown) (**A**) and quantification of CTSO positive area (**B**) in human unruptured and ruptured IA domes. The black rectangles represent the region shown at higher magnification in the bottom right-hand corner of each image. (Results were expressed as mean ± SEM; \**p* < 0.05; Mann-Whitney test).

### *Ctso* silencing does not affect VSMC proliferation and apoptosis

We next aimed to elucidate the role of CTSO in the arterial wall by focusing on VSMC, in which it is predominantly expressed and its secretion is modulated by stretch. To this end, we evaluated the consequences of siRNA-mediated knockdown of endogenous CTSO in VSMC (KD-SMC) in comparison with a control siRNA pool (Cont-SMC). *Ctso* silencing did not affect VSMC proliferation (Figure 4A), nor apoptosis as measured by TUNEL and caspase 3 cleavage in basal condition (no apoptosis, data not shown) and after induction of apoptosis by staurosporine (Figure 4B and C). These results indicate that CTSO did not affect VSMC proliferation and viability.

**Figure 4.**
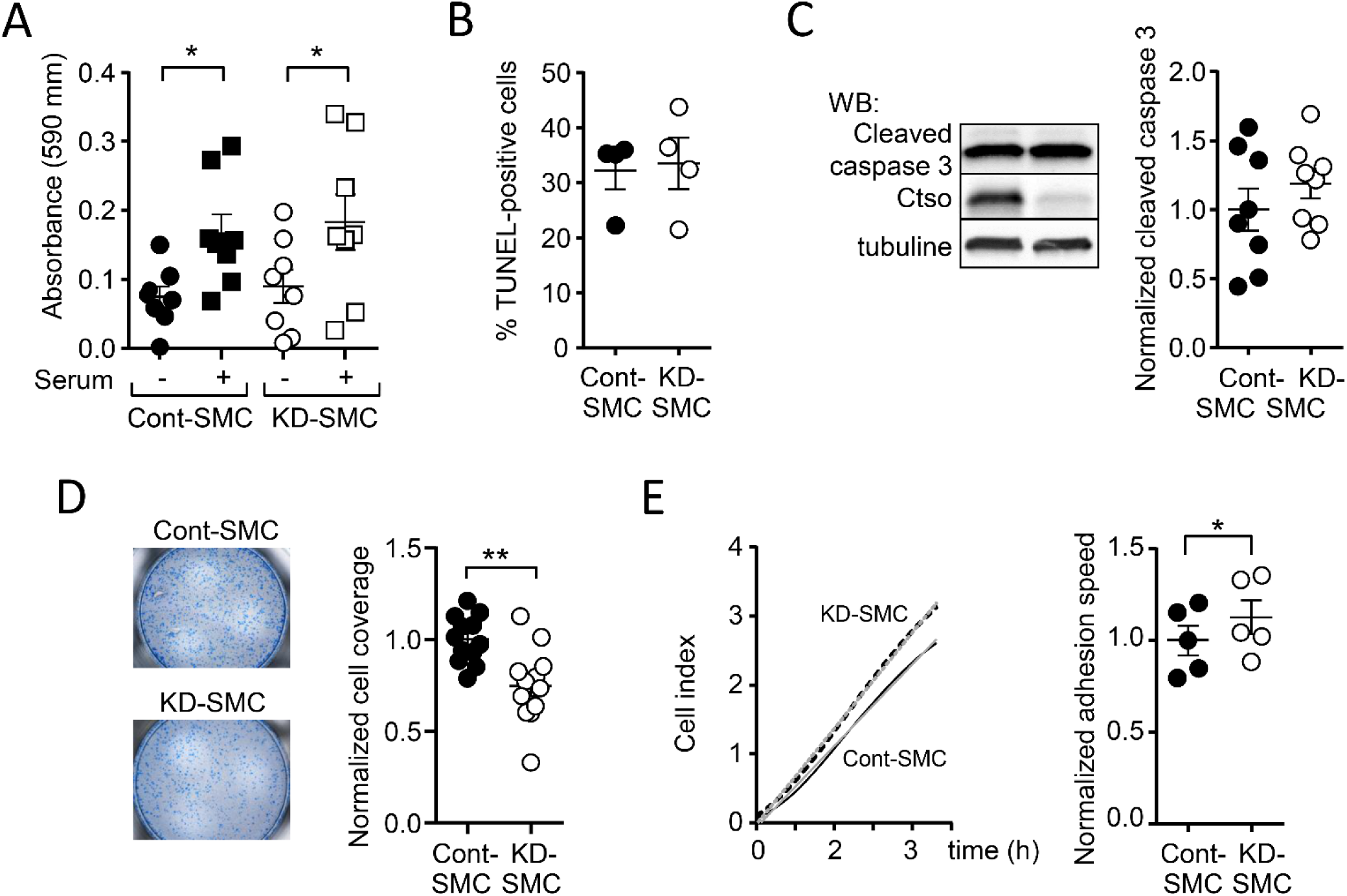
Consequence of CTSO depletion on VSMC proliferation, apoptosis, migration and adhesion. **(A**) Proliferation assessed by the measurement of MTT absorbance in the absence and presence of 10% serum in VSMC transfected with siRNA targeting *Ctso* (KD-SMC) and control siRNA (Cont-SMC). **(B**), Number of apoptic cells in KD-SMC and Cont-SMC culture after stimulation with staurospaurine (1 µmol/L, 8 h). Values represent the percentage of TUNEL-positive cells relative to total cell population in each group. **(C**) Typical Western blot of cleaved-caspase 3 in Cont- and KD-SMC and corresponding quantification. CTSO and tubulin have been also blotted to check CTSO silencing and equal loading. **(D**) Representative images of Cont- and KD-SMC which migrated to the underside surface of transwell membranes and the corresponding quantification. **(E**) Representative curve showing the cell index representing the adhesion of Cont- and KD-SMC over time and quantification of the adhesion speed. (Results were expressed relative to Cont-SMC set as 1 (except in B). Data are presented as mean ± SEM; \**p* < 0.05, \*\**p* < 0.01; one-way *ANOVA* in A; Mann-Whitney test in (B-E)).

### *Ctso* silencing modifies VSMC migration and adhesion

We next tested whether CTSO regulates the migratory and adhesive properties of VSMC using Boyden chambers and adhesion assays, respectively. CTSO depletion in VSMC led to a significant decrease in transmigration (Figure 4D), but accelerated the adhesion speed of VSMC on fibronectin (FN) matrix (Figure 4E). This effect was associated with an increased phosphorylation of focal adhesion kinase (FAK) in KD-SMC compared to cont-SMC, both at 2 h and 24 h after seeding on FN matrix (Figure 5A). When VSMC were plated on plastic, without FN coating, a significant increase in FAK phosphorylation was also observed at 24 h but not at 2 h post-seeding (Figure 5B). It is known that VSMC produce and release FN which, in turn, can modulate their phenotype and functions (19). The observed differences in FAK phosphorylation induced by CTSO depletion may therefore results from an effect on the endogenous FN. To test this hypothesis, we thus analyzed the effect of *Ctso* silencing on FN expression in Cont- and KD-SMC cultures. Western blot analysis revealed an increased level of FN in KD-SMC compared to Cont-SMC, while there was no difference in collagen I (COL-I) expression (Figure 5C). This rise in FN expression in KD-SMC was not associated with a change in mRNA-*Fn1* level which, like mRNA-*Col1a*, remains the same in Cont- and KD-SMC (Figure5D). The observed increase in FN level induced by CTSO silencing could therefore stem from the ability of CTSO to degrade FN. To test this hypothesis, we cultured Cont- and KD-SMC on human FN, which we quantified by immunofluorescent labeling. FN labelling confirmed the presence of a greater amount of FN in the vicinity of KD-SMC compared to Cont-SMC (Figure 5E). All together, these data show that CTSO controls VSMC migration and adhesion, and suggest that it could result from an effect on FN.

**Figure 5.**
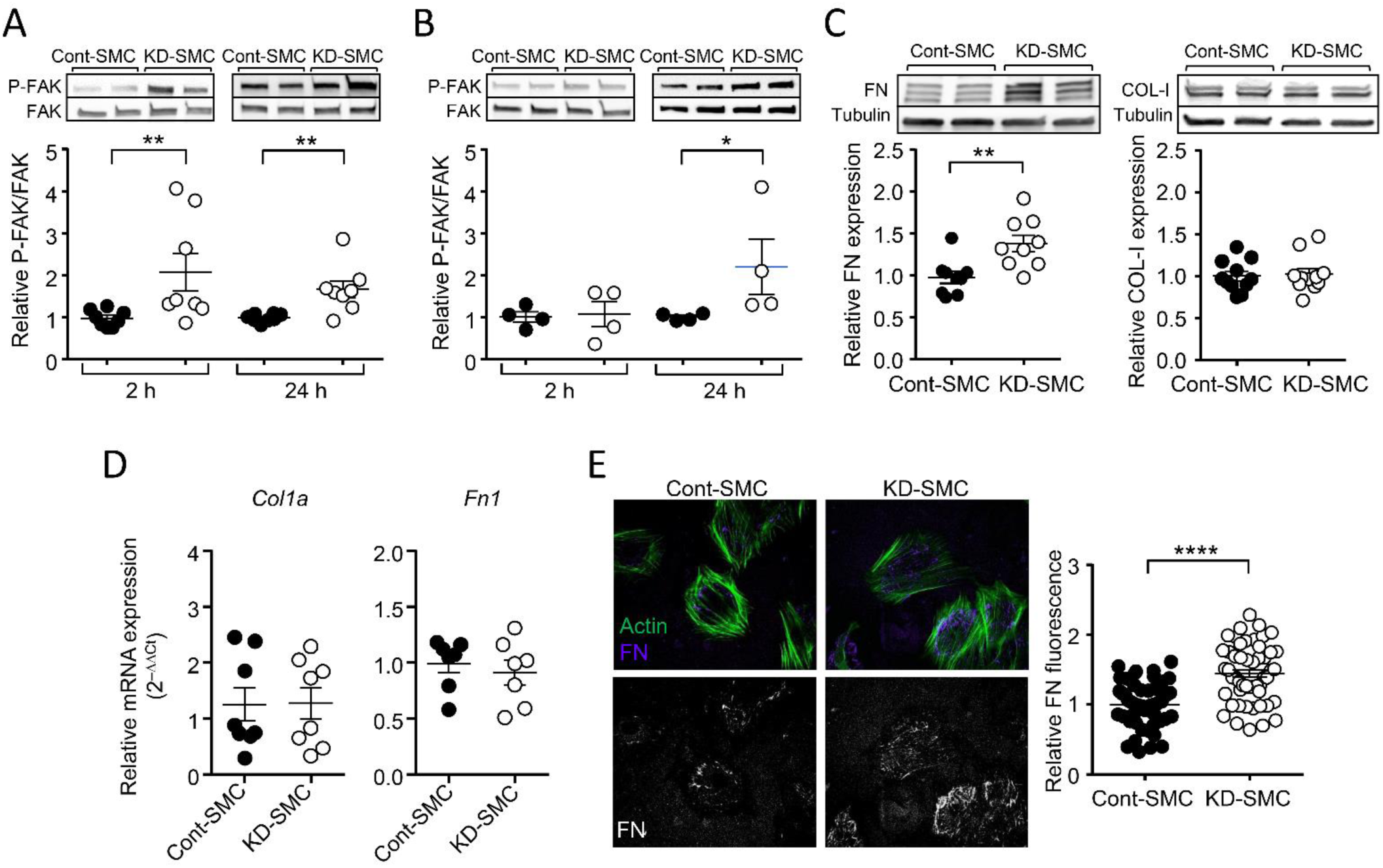
CTSO depletion in VSMC stimulates FAK phosphorylation and FN expression. (**A** and **B)** Representative Western blots showing the phosphorylation of FAK in Cont- and KD-SMC 2 h and 24 h after seeding on FN matrix (**A**) or directly on plastic (**B**). Graphs show densitometric analysis of phospho-FAK in blots, relative to FAK expression. **(C**) Representative western blots showing the amount of FN and Col-I in Cont- and KD-SMC and corresponding quantification. **(D**) RT-qPCR analysis of mRNA levels of Col1A, Fn1. Results were expressed relative to Cont-SMC and expressed as mean ± SEM (ns, Mann-Whitney test). **(E**) Representative immunofluorescent staining of human FN (purple) and phalloidin staining of actin fibers (green) in Cont- and KD-SMC (top images). Isolated FN staining is also shown (white, bottom images). SMC were seeded on human FN-coated surface. Quantification of FN has been made cell by cell (each dot represents a cell, n=4 independent experiments). (Results were expressed relative to Cont-SMC and presented as mean ± SEM; \**p* < 0.05, \*\**p* < 0.01, \*\*\*\**p* < 0.0001; Mann-Whitney test).

### *Ctso* silencing affects contractile phenotype marker gene expression

We next assessed the potential role of CTSO on VSMC phenotype by measuring the expression of contractile marker gene expression (Figure 6A). Compared with Cont-SMC, the expression of all tested VSMC marker mRNAs, namely *Acta2, Myh11*, *Tagln* and *Cnn1* was significantly up-regulated in KD-SMC, which showed a 90%-decrease in *Ctso* mRNA expression (Figure 6B). By contrast, the expression of *Bmp4*, a known marker of osteogenic differentiation of VSMC (20) was reduced in KD-SMC compared to Cont-SMC (Figure 6B). These results suggest that *Ctso* silencing reinforced VSMC contractile phenotype. However, measurement of contractility showed that KD-SMC contracted collagen gel less strongly than Cont-SMC in response to the thromboxane A2 analog U46619 (Figure 6C).

**Figure 6.**
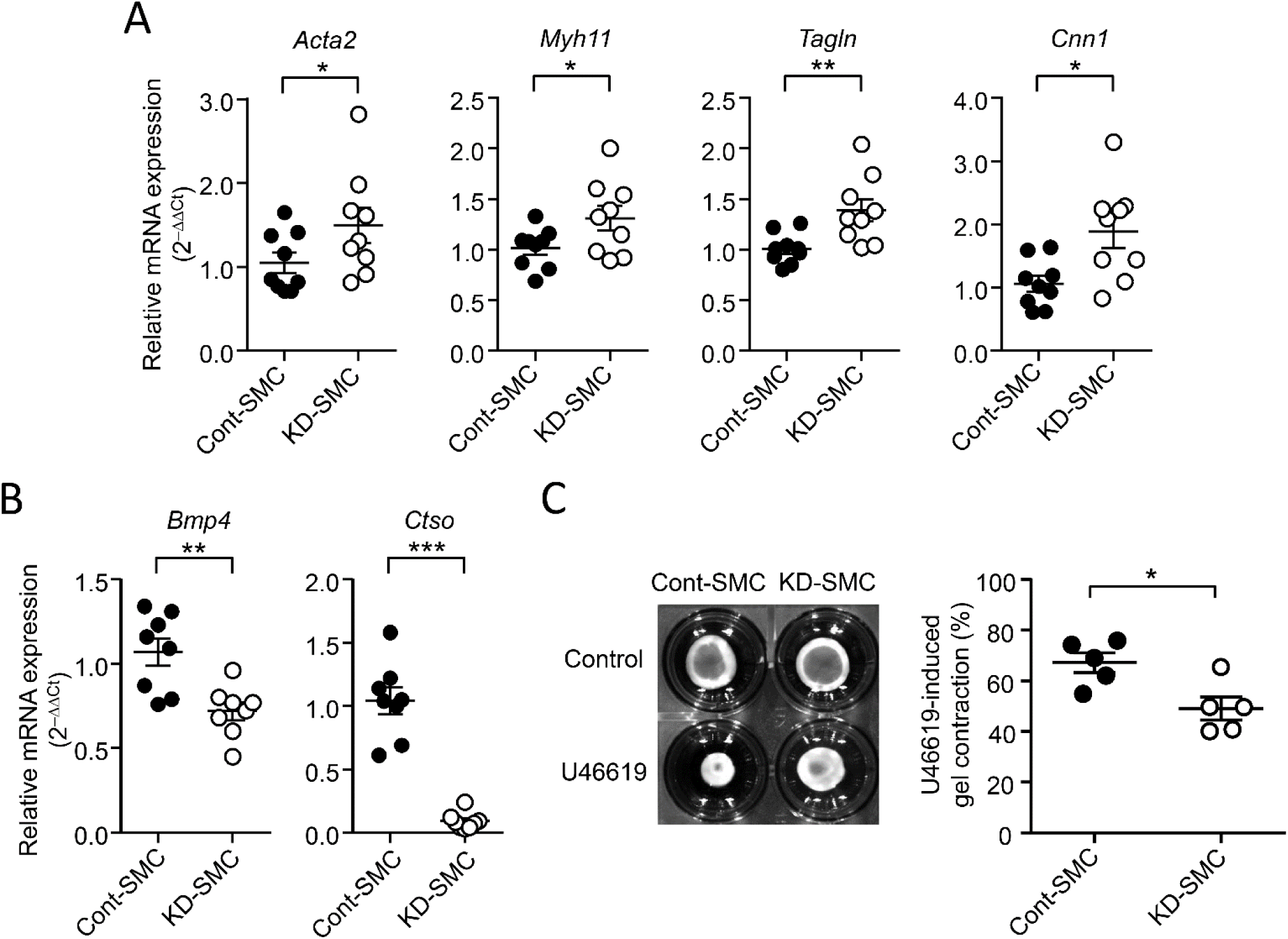
CTSO depletion in VSMC modulates contractile phenotype marker expression and contraction. (**A-B**), RT-qPCR analysis of mRNA levels of the SMC marker genes Acta2, Myh11, Talgn, Cnn1 (**A**); and Bmp4 and Ctso (**B**) in Cont- and KD-SMC. Results were expressed relative to Cont-SMC and expressed as mean ± SEM; *p < 0.05, **p < 0.01, ***p < 0.001; Mann-Whitney test). (**C**) Representative images of Cont-SMC- and KD-SMC-seeded collagen gels 48 h after release with and without U46619 and quantification of U46619-induced gel contraction in Cont- and KD-SMC. (Data are expressed as mean ± SEM; \**p* < 0.05; Mann-Whitney test).

### *Ctso* silencing induces stiffening of VSMC

We then used atomic force microscopy (AFM) to assess the effect of CTSO depletion on VSMC and VSCM-derived ECM stiffness, as arterial stiffening is known to be a risk factor for IA (21). Elastic modulus determined by AFM nanoindentation showed that the stiffness of KD-SMC was significantly higher than that of Cont-SMC, and was strongly reduced by complementation with exogenous CTSO (Figure 7A and S2). In contrast, the corresponding ECM produced by KD- and Cont-SMC displayed a similar stiffness (Figure 7A and S2).

**Figure 7.**
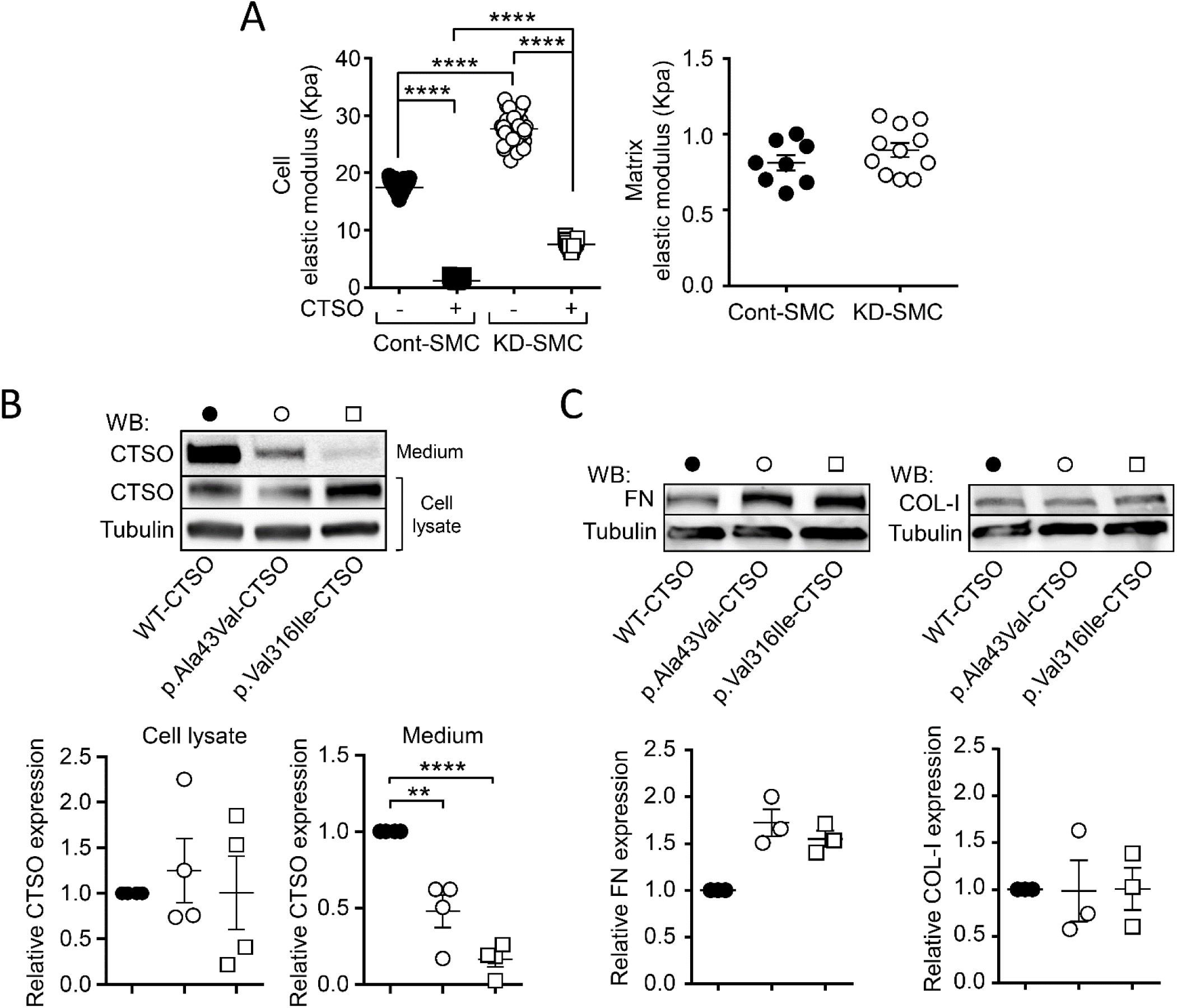
Effect of CTSO depletion and CTSO variant expression on cell stiffness and FN expression. (**A**) Atomic force microscopy determination of the elastic modulus of Cont- and KD-SMC, in the absence and presence of exogenous CTSO (200 ng/ml) (left), and their respective decellularized extracellular matrix (right). (Data are expressed as mean ± SEM; \*\*\*\**p* < 0.0001; one-way *ANOVA*). (**B**) Representative western blots and quantification of CTSO protein expression measured in cell lysate and culture medium of NIH3T3 expressing WT-CTSO, p.Ala43Val-CTSO and p.Val316Ile-CTSO. (**C**) Representative western blots and quantification of FN and Col-1 expression measured in WT-CTSO, p.Ala43Val-CTSO and p.Val316Ile-CTSO. (Data are expressed as mean ± SEM; \*\**p*<0.01 and \*\*\*\**p* < 0.0001; one-way *ANOVA*)

### CTSO variants

To assess the impact of c.128C>T *CTSO* and c.946G>A *CTSO* variants on CTSO protein we established stable cell lines expressing wild-type (WT-CSTO), and the two mutated proteins, p.Val316Ile-CTSO and p.Ala43Val-CTSO. Western blot analysis showed that expression level in cell lysates was similar for WT-CTSO and the two mutated CTSO proteins (Figure 7B). In contrast, the amount of p.Val316Ile-CTSO and p.Ala43Val-CTSO in the culture medium was strongly reduced compared to WT-CTSO (Figure 7B). Since we observed an increase in the amount of FN after CTSO depletion in VSMC (Figure 5C), we investigated the effect of p.Val316Ile-CTSO and p.Ala43Val-CTSO expression on FN level. Western-blot results show that the expression of CTSO mutants mimicked the effect of CTSO depletion on the increase of FN level, without affecting COL-I expression (Figure 7C).

## Discussion

We have identified two rare missense variants in the *CTSO* gene shared by all the affected relatives in two large pedigrees with multiple IA-affected relative. Of note, we also observed several subjects (n=8) carrying possibly deleterious *CTSO* variations but without IA at the time of the study. In addition, we found a significant increase in the proportion of subjects with history of high blood pressure among IA carriers compared to unaffected individuals. High blood pressure history is a well-established environmental factor associated with the development of IA (22). Our results suggest, as described previously in familial cases with rare coding variants in *ANGPTL6*,(16) that rare coding variants in *CTSO* could promote IA formation in combination with other deleterious genetic or environmental factors such as high blood pressure, which would act either directly by mechanical effects on the vessel walls or indirectly by triggering inflammation (23).

CTSO is one of the 11 cathepsins encoded in the human genome which constitute an important group of proteases that regulate numerous processes (18). Cathepsins are highly expressed in intracellular acidic compartments such as endosomes and lysosomes. Loss of function mutations in their encoding genes cause very different syndromes in terms of clinical symptoms and disease progression, corresponding to either typical lysosomal storage diseases, or resulting from defective cleavage of specific protein substrates (24). Cathepsins are also found in the cytoplasm, cell nucleus, cell membrane and the extracellular space (25, 26). Extracellular cathepsins mediate ECM protein degradation (collagen, elastin, fibronectin, laminin) and the release of ECM-bound factors, but also shed various membrane proteins including receptors, growth factors, cytokines, and adhesion proteins thereby influencing important cellular processes such as proliferation and differentiation, motility, cell-cell interaction, adhesion, inflammatory and immune responses. Indeed, extracellular cathepsins are majorly upregulated in pathological states and are implicated in a wide range of disorders including vascular remodeling and atherosclerotic diseases (27, 28). The cathepsins B, K and S have been found to be upregulated in the wall of IA (29). Here we observed an increase in CTSO expression, in particular in ECM, in ruptured human IA compared to unruptured IA. At cell level, CTSO is expressed in endothelial cells, and more strongly in VSMC. CTSO was found both in and around VSMC, and its extracellular release is stimulated by cell stretching, suggesting that its role as an extracellular protease in the arterial wall could be potentiated by high blood pressure and participated to the adaptive arterial remodeling induced by hypertension. The increase in CTSO expression observed in ruptured IA domes could thus result from the greater aneurysmal wall stretch induced by blood pressure in ruptured *versus* unruptured IA (30).

CTSO is defined as a ubiquitous protein but neither its peptidase activity nor its function had been characterized yet. Although we have not analyzed the enzymatic activity of CTSO *in vitro* to directly demonstrate its ability to degrade FN, we provide a strong body of experimental evidence to support this hypothesis. Cell adhesion and FAK activation are potentiated by depletion of CTSO, both rapidly and late after VSMC seeding on exogenous FN coating, while only delayed stimulation of FAK activation was observed in CTSO-depleted VSMC seeded on plastic. These observations are consistent with a degradation of pericellular FN by CTSO, which instantaneously impaired cell-FN coating interaction in VSMC producing extracellular CTSO. When VSCM are seeded on plastic, the effect of CTSO depletion is only visible at late time points, after the time necessary for the cells to self-produce FN with which they establish adhesions. This hypothesis is in agreement with the increase in the amount of FN in CTSO deficient VSMC and is further supported by the strong immunofluorescence labeling of exogenous FN close to CTSO-depleted VSMC compared to its low intensity in CTSO-expressing cells. All these observations suggest that extracellular CTSO secreted by VSMC degrades FN. This was corroborated by the results obtained by the expression of the two mutated proteins p.Val316Ile-CTSO and p.Ala43Val-CTSO which, being weakly secreted, induce an increase in the amount of FN as does the depletion of CTSO.

It is well established that adhesive properties to ECM drives VSMC stiffness (31), and the level of activation of FAK signaling is related to VSMC stiffness (32). In particular, VSMC focal adhesions to FN correlate with VSMC stiffness (33). The strong stiffening of VSMC that we observed after CTSO depletion is therefore consistent with an increase in the ability of VSMC to form adhesions to FN due to the reduced degradation of FN in CTSO-depleted VSMC.

ECM composition and organization impact the physical interactions with VSMC, which play a major role in regulating their functions, including migration and differentiation. We observed that knocking-down CTSO reduced VSMC migration on FN. This result further supports a role of CTSO in promoting FN degradation, although additional mechanisms could also be involved such as degradation of other extracellular proteins or lysosomal degradation of FN-bound integrins known to be required for migration.(34) Regarding VSMC differentiation, CSTO depletion leads to increased expression of contractile phenotype marker genes such as *Acta2*. Interestingly, deletion or inhibition of cathepsin B, characterized as a FN-degrading protease (35) that is also up-regulated by cell stretching, also increases ACTA2 expression through an underlying mechanism that is still unknown (36). Increased ACTA2 expression has been shown to contribute to VSMC stiffening (37), suggesting that up-regulation of *Acta2* and possibly of the other contractile markers, could participate to the increase in VSMC stiffness induced by CTSO depletion. Although contractile markers were upregulated, collagen gel contraction produced by KD-VSMC was reduced compared to Cont-VSMC. Whether this effect was related to the stiffness of VSMC induced by CTSO deletion could not be determined. Nevertheless, this possibility is supported by previous results obtained on VSMC from aged animals showing a similar association between increased stiffness (31) and reduced collagen gel contraction (33, 38).

Besides their contractile activity, VSMC produce ECM that allows adaptation to mechanical forces that act on the vessel wall while maintaining adequate wall pressure (39, 40). Thus, the composition, organization and resulting interactions of ECM with VSMC are adjusted to the mechanical demands of the vessel wall through multiple and coordinated mechanisms. Among them, the fine-tuning of extracellular protease expression and activity plays a major role in ECM matrix reshaping (27). Our present study proposes that CTSO, whose secretion is regulated by cell stretch, could be a new member of these ECM proteases involved in arterial wall adaptation to mechanical stress. Indeed, we have identified CTSO as a potential new player in arterial remodeling, regulating FN deposition and VSMC function. This role of CTSO is consistent with an increased susceptibility to IA associated with non-secreted c.128C>T *CTSO* and c.946G>A *CTSO* variants. We suggest that, by promoting increased FN deposition, VSMC-FN adhesion and VSMC stiffening, the p.Val316Ile-CTSO and p.Ala43Val-CTSO mutants affect the mechanical properties of the vessel wall and compromise the proper adaptation of the arterial wall to local hemodynamics, thereby favoring IA formation. It is noteworthy that similar features, namely faster adhesion to FN,(41) up-regulation of contractile marker expression and increased stiffness, have been described in Marfan syndrome VSMC,(42) a well-known genetic condition that predisposes to IA (43).

## Methods

### Sex as a biological variable

For experiments in SHR and SHRSP, only males have been used as males showed higher blood pressure and greater incidence of stroke than females (44). For analysis in humans, both males and females have been included.

### Clinical recruitment

Familial cases of IA are defined as at least two first-degree relatives both diagnosed with typical IA (defined as a saccular arterial dilatation of any size occurring at a bifurcation of the intracranial vasculature), without any age limitation. Index case subjects and their relatives were recruited according to a recruitment process previously described (45). Briefly, neuroradiological phenotyping was performed in each recruiting center by interventional neuroradiologists, neurologists, and neurosurgeons in order to recruit only case subjects with typical saccular bifurcation IA. Mycotic, fusiform-shaped, or dissecting IAs were systematically excluded, as well as IA in relation with an arteriovenous malformation and IA resulting from syndromic disorders such as Marfan disease or vascular forms of Elhers Danlos. For all included patients we recorded the date of birth, if IA, the cases of rupture, the number of IA, the IA larger diameter in mm, the IA locations according to the four following groups internal carotid artery (ICA), posterior cerebral circulation (PCA), middle cerebral artery (MCA) or anterior cerebral artery (ACA), the smoking habits, the history of high blood pressure and ischemic stroke, the level of alcohol intake per week, the history of diabetes or dyslipidemia, obesity (BMI >30), treatment by statin, antiplatelet, oral anticoagulant and anti-inflammatory drugs.

### Whole Exome Sequencing (WES)

Genomic DNA was extracted from peripheral blood lymphocytes using the NucleoSpin Blood kit XL (Macherey Nagel). Whole-exome fragments were captured with the SureSelect Human All Exon V4 kit (Agilent technology), according to manufacturer protocol. Alignment on reference genome (Broad Institute human_g1k_v37) was performed using Burrow-Wheeler aligner (Bwa mem v0.7.10).(46) Picard v1.119 was used to flag duplicates and recalibration was achieved through GATK v3.2.2 (broadinstitute, 2023) (47). Variants were called with GATK HaplotypeCaller on all exons hg9 exons (NCBI RefSeq), annotated with SnpEff and vcfGnomad (jvarkit) using gnomad v2.1.(48) Quality filters detailed in the Table S1 were applied. Only variants annotated as non-synonymous or affecting splicing, and reported with a minor allelic frequency (MAF) below 0.1% in the non-Finnish European (NFE) gnomAD population, were subsequently considered.

### IBD analysis

Haplotype sharing between affected relatives were identified through IBD analysis based on SNP genotyping data. Fluorescence intensities were obtained from Precision Medicine Research Array (PMRA - Affymetrix) and quantified by Affymetrix GeneTitan Multi-Channel Instrument. Genotypes of affected relatives II.2, II9, III.2, III.4 and III.7 were merged with PREGO control population (49) and SNPs with MAF<10%, call rate < 95% or p<1.10-5 when testing for Hardy-Weinberg equilibrium were excluded. IBD regions were then identified using IBDLD v3.34 with noLD method (50). Familial segregation analysis by capillary sequencing was performed on an Applied Biosystems 3730 DNA Analyzer, using standard procedures. Sequence analyses were performed with SeqScape v.2.5.

### Human Saccular IA Samples

Saccular IA samples are from the Aneux Biobank (51). All samples were obtained during microsurgery by resection of the IA dome (i.e., the bulging region of the IA) after clipping of the neck performed at the Division of Neurosurgery of the Geneva University Hospitals, Switzerland. IAs were stored as previously described (51), fixed in formol, embedded in paraffin, sectioned at 5 μm and conserved at 4°C.

### Immunohistochemistry

Labeling of 4-μm-thick sections was performed following heat-induced epitope retrieval in citrate buffer (10 mM, pH 6.0). Sections of unruptured (n=10) and ruptured (n=10) IA were immunolabeled with the Prestige Antibodies® antibody recognizing CTSO (HPA002041, Sigma-Aldrich). Negative controls were performed without CTSO antibody. Hematoxylin and eosin were used as counterstaining. Image acquisition of immunohistochemical staining of CTSO was done with a whole-slide scanner NanoZoomer (Hamamatsu). Total tissue area and positive staining area were quantified using Image-Pro Plus (Media Cybernetics), and the positive CTSO staining was expressed relative to total sample area.

### Animal model

Cerebral arteries from 3 months-old Wistar-Kyoto (WKY), spontaneously hypertensive (SHR) and stroke-prone SHR (SPSHR) male rats (300 g) were collected and snap-frozen in liquid nitrogen for RNA extraction. WKY were used for VSMC isolation.

### Cell culture

Primary VSMC were isolated from the aorta of 4-week-old WYK rats. Rats were sacrificed according to institutional animal handling ethics. The aorta was harvested, cleaned, endothelium-denuded, cut into small pieces and digested by collagenase II (1 mg/mL; 2 h at 37°C under agitation; Worthington Bio-chemical). After inactivation of collagenase with serum, the tissue was spun down and plated in a 6-well plate in Dulbecco modified Eagle medium (DMEM, Gibco; Invitrogen) containing 10% foetal bovine serum (FBS), 4.5 g/L glucose, 100 units/mL penicillin and 100 µg/mL streptomycin at 37°C-5% CO2. All experiments were performed at passages 2 and 3. For cell stretching, cyclic uniaxial strain was applied at 1 Hz and 10% or 20% strain for 24 hours (MechanoCulture FX, CellScale). HUVEC (passage 2 to 6; PromoCell) were routinely cultured in EBM media supplemented with the provided growth factors kit (Promocell). For flow experiments HUVEC were cultured on 0.2% gelatin-coated 0.4 ibidi slides (IBIDI) and unidirectional laminar shear stress was applied using the pumping system and control unit form IBIDI. Local shear stress was calculated using Poiseuille’s law and averaged to 3 dyn/cm^2^ (pathological low shear stress: LSS) 16 dyn/cm^2^ (physiological shear stress: PSS) or 36 dyn/cm^2^ (pathological high shear stress: HSS). NIH3T3 cells were cultured in DMEM containing 1 g/L glucose, 10% FBS, 100 units/mL penicillin and 100 µg/mL streptomycin at 37°C-5% CO2.

### RT-qPCR

Total RNA was purified from cells using RNA plus kit (Macherey Nagel) or tissue using TRIzol reagent (MAN0001271, Ambion, Thermo Fisher Scientific) and RNA XS Plus kit (Macherey Nagel) according to the manufacturer instructions. RNA (500 ng) was reverse-transcribed with M-MLV enzyme (28025021, Thermo Fisher Scientific). Real-time qPCR was performed on a 7900HT Fast Real-Time PCR System (Applied Biosystems) using SYBR Green Master Mix (4367659, Applied Biosystems) and primers listed in Table S4. Each sample was analyzed in triplicate. GAPDH was used as the reference gene and results are expressed according to the 2^-ΔΔCt^ method.

### Immunoblotting

Supernatant from VSMC or NIH3T3 were taken off and concentrated with Amicon Ultra-0.5 Centrifugal Filter Unit (UFC5010, Millipore). Cells were lysed on ice in buffer supplemented with protease and phosphatase inhibitor cocktails (Sigma Aldrich) and sodium orthovanadate. Equal amount of proteins of each sample was separated by SDS-PAGE, transferred to nitrocellulose membranes, and incubated with specific antibodies: CTSO (for rat form: ab200735, Abcam, for human form: HPA002041, Sigma-Aldrich), cleaved caspase 3 (9664, Cell Signalling Technology), P-FAK (8556, Cell Signalling Technology), FAK (3285, Cell Signalling Technology), Fibronectin (15613-1-AP, Proteintech), type I collagen (14695-1-AP, Proteintech). Equal loading was checked by reprobing of the membrane with an anti-tubulin antibody (T9026, Sigma). Immune complexes were detected with appropriate secondary antibodies and enhanced chemiluminescence reagent (Clarity ECL BioRad). Protein band intensities were quantified using ImageJ Software (NIH software, Bethesda, Md).

### Ctso silencing

VSMC were transfected using Lipofectamine RNAiMAX reagent (Invitrogen) according to the manufacturer instructions with a pool of siRNAs targeting rat *Ctso* (ON-TARGETplus siRNA-SMARTpool (Cat# L-110260-00-0005; Horizon Discovery,) and a non-targeting control pool (Pool #1, D-001810-10-05, Horizon Discovery) to generate KD-SMC and Cont-SMC, respectively. The efficiency of *Ctso* mRNA depletion was assessed 72 h post-transfection by RT-qPCR.

### Viability/Proliferation assay

VSMC viability/proliferation was quantified by MTT (3-[4,5 Dimethylthiazol-2-yl]-2,5-diphenyltetrazodium) colorimetric assay according to the manufacturer instructions (M5655; Sigma-Aldrich). Briefly, VSMC were seeded onto 96-well plates in triplicate for each condition, cultured for 24 h, after which they were serum-starved for 24 h. VSMC were then incubated in DMEM without or with 10% FBS for 24h. The number of viable cells was then determined by incubation in 1 mg/mL MTT for 4 h à 37 °C. The medium was then removed and acidified isopropanol was added to solubilize the MTT reduction product formazan. MTT reduction by viable cells was then quantified by measurement of the absorbance at 590 nm.

### Apoptosis assay

VSMC apoptosis was determined after incubation in 1 µmol/L staurosporine (S5921, Sigma-Aldrich) in serum-free medium for 8 h. Apoptosis was quantified by the measurement of cleaved caspase 3 by western blot and by TUNEL assay. VSMC were fixed with 4% paraformaldehyde and permeabilized. Fragmented DNA was stained with TUNEL Assay Kit - BrdU-Red (ab66110, Abcam) according to manufacturer instructions, and DAPI was used to label all nuclei. Coverslips were mounted on slides with Prolong gold antifade reagent and observed with a fluorescent microscope. Apoptotic cell number was quantified as the percentage of TUNEL-positive cells.

### Transwell migration assay

Boyden chamber assay was performed as previously described (52). VSMC, transfected with siRNA 48 h before, were trypsinized and washed in serum-free DMEM before plating into transwells (0.47cm^2^ of culture area, 8 µm pore size; Nunc™). DMEM containing 10% FCS was placed in the lower chamber and VSMC were allowed to migrate for 10 h at 37 °C. After incubation, the filter was removed and VSMC on the upper side of the filter were scraped off. VSMC that had migrated to the lower side of the filter were fixed with 4% paraformaldehyde and stained with Coomassie blue 0.1% for observation under a microscope in brightfield mode. Migration to the lower chamber of the transwell was quantified by the area covered by cells measured using ImageJ Software and expressed expressed relative to Cont-SMC set as 1.

### Cell adhesion assay using impedance technology

VSMC (10000/well) were seeded in a 96 well plate microtiter xCELLigence assay plate coated with 2 µg/ml fibronectin (E-Plate, ACEA Biosciences Inc.) and placed on the Real-time xCELLigence Cell Analyzer (Roche Applied Science) platform at 37°C. The cell index value, which are proportionate to the area covered by the cells was measured every 5 min for a period of 8 h. Adhesion speed was defined as the slope of the cell index change over time.

### Collagen gel contraction assay

VSMC were mixed with collagen gel working solution (CBA-201, Cell Biolabs). The cell-collagen mixture was added into a 24-well plate and incubated at 37°C for 1 h to allow collagen polymerization. Serum-free DMEM was added to the top of the collagen gel lattice. After 24 h, cells were treated or not with U46619 (10^-6^ mol/L) and the myosin inhibitor BDM (from CBA-201 kit), and the collagen gels were released by a sterile spatula. Changes in the collagen gel area were measured at 48 h. The percentage of contraction corresponded to [(area in BDM condition-area in the control or U46619 condition)/ area in BDM condition × 100].

### Immunofluorescence

VSMC were seeded onto culture plates pre-coated with human FN (F2006, Sigma-Aldrich, 20 µg/ml). After 24-hour of culture, cells were fixed with 4% paraformaldehyde for 15 min, rinsed twice with PBS, then permeabilized and blocked for 1 h with carbonate-bicarbonate buffer containing 1% FBS, 3% BSA, 0.5% Triton X-100, 0.01% deoxycholate, 0.02% sodium azide. FN was stained with anti-human FN antibody (F3648, Sigma-Aldrich; 1/500 overnight), revealed by a secondary Alexa647-conjugated donkey anti-rabbit antibody (LifeTech; 1/1000, 2 h). Actin staining was performed at room temperature using phalloidin Alexa Fluor™ 488 (A12379, Fisher Scientific; 1/400, 45 min). Images were taken using Nikon A1 confocal microscope (Nikon France, Champigny sur Marne) equipped with a x60 Plan-Apochromat objective with a numerical Aperture of 1.4. FN fluorescence was quantified using FIJI. Briefly, for each cell a mask corresponding to the cell area was generated and applied on the FN staining image. FN fluorescence intensity in this area was extracted for each cell.

### Atomic force microscopy (AFM) experiments

AFM experiments have been performed in Cont- and KD-SMC grown on plastic and allowed to produce their own ECM for 5 days. ECM analysis has been performed after decellularization as previously described (53). All experiments were then performed in Physiological Saline Solutions (PSS) buffered with 20 mM HEPES at 37°C using a NanoWizard® atomic force microscope (JPK Instruments, Germany) equipped with a temperature controller and inverted optical microscope. Indentation experiments were carried out with cantilevers (SQube) having a colloidal glass sphere of 5 µm in diameter. Their spring constant was calibrated using the thermal noise method implemented in the AFM setting (JPK software), with values comprised between 0.12 and 0.15 N.m^-1^. Prior to indentation measurements, the cantilever sensitivity was systematically measured from the slope of force-distance curves performed on glass. The optical microscope was first used to position the AFM tip on ECM or on the cytoplasmic region of the selected cell and several approach-retract force-distance curves were performed to determine the force to be applied corresponding to an indentation depth of ∼500 nm (a good compromise avoiding the long-range interactions and in the validity domain of the Hertz contact mechanics model). Then, force-distance curves were recorded on cells at a low constant speed of 1 µm/s to neglect the hydrodynamic drag forces exerted on the cantilever by the liquid medium (54). These curves were then converted into force-indentation curves, and the approach part was fitted by the Hertz model to determine the apparent elastic modulus of cells (55).

### Mutagenesis

CTSO-TurboGFPpPlasmid was purchased from Origene (RG208268). c.128C>T *CTSO* and c.946G>A *CTSO* mutagenesis was performed using Q5® Site-Directed Mutagenesis Kit Protocol (E0554, NEB) and designed primers: A43V-Fw: 5’-GAA GCCGCCGTCTTCCGGGAAAGTC-3’; A43V-Rv: 5’-ACG CTCGCGGCTCCGCGG-3’; V316I-Fw: 5’-GTTTGTGGTATTGCAGATTCCATTTCTTCTATATTTGTGACGC-3’; V316I-Rv: 5’-GCGTCACAAATATAGAAGAAATGGAATCTGCAATACCACAAAC-3’. Mutations were confirmed by standard sequencing methods using T7 standard primer and designed primer (5’-AAGCCCCTGGAAGACCTAAG-3’).

### Generation of lentiviral constructs

From LT3GEPIR plasmid (#111177, Addgene), was modified to remove miR30 backbone by amplifying and cloning eGFP between XhoI and EcoRI restriction sites using the following primers: Fw 5’-CGGCCGCTCGAGATGGTGAGCAAGGGCGAGGAG-3’and Rv 5’ GATCTGAATTCTTACTTGTACAGCTCGTCCATGC-3’. The resulting plasmid was used to clone WT and mutant CTSO in BamHI and XhoI,

### Lentivirus production, transduction and induction of WT-CTSO and CTSO variants

Lentiviral vector for expressing WT-CTSO, p.Ala43Val-CTSO and p.Val316Ile-CTSO was transfected into HEK293T cells along with packaging vectors psPAX2 and pVSVG2 (provided by Dr. Utz Fischer’s lab) using polyethylenimine (#764965, Sigma-Aldrich) transfection agent. After overnight incubation, the medium was replaced with fresh medium (DMEM+10% serum). Forty-eight hours and 72 h after transfection, viral supernatant was collected, filtered through 0.22 µm filter and used to infect NIH3T3 in the presence of polybrene (8 µg/mL, H9268; Sigma-Aldrich). After 48 to 72 h of infection, transduced NIH3T3 cells were selected with puromycin (2 µg/mL, P8833, Sigma-Aldrich) for 48 h and maintained in the presence of puromycin (2 µg/mL). The induction of WT- or mutated CTSO was performed at least 2 days before the experiment by supplementing the medium with doxycycline (1 µg/mL, #D9891, Sigma Aldrich), which was maintained until the completion of the experimental procedure.

### Statistical analysis

Data are expressed as the mean ± SEM of sample size n. All values use biological replicates and are indicated by group size n in figure legends or within graphs. For *in vivo* data, each n value corresponds to a single animal. For *in vitro* data, each n value corresponds to an independent experiment. Investigators were blinded for some measurements made (i.e., immunofluorescence, immunohistology, area measurement). Comparisons between two groups were performed by unpaired, two tailed nonparametric Mann-Whitney *U* test, and one-way ANOVA with relevant *post hoc* tests was used for multiple-group comparisons using GraphPad Prism 6.0 software (GraphPad Software). *P* < 0.05 was considered statistically significant.

### Study approval

All animal care and use procedures of the present study were performed in accordance with the European Union Directive 2010/63/EU about the protection of animals used for scientific purposes and were authorized by the Ethics Committee on Animal Experimentation from the Région Pays de la Loire (approval number D44278). For genetic analysis, index case subjects with IA and their relatives were recruited following the French ethical guidelines for genetic research and under approval from the French Ministry of Research and the local ethical committee (no. DC-2011-1399; Clinical Trial NCT02712892). Informed written consent was obtained from each individual agreeing to participate in the genetic study, to whom MRI screening and blood sampling were proposed. AneuX biobank was approved by the Geneva State Ethics Commission for Research involving humans (Geneva CCER 2022-00426). All patients approved for the use of their data and biological samples in the field of cerebrovascular research. Research was conducted in accordance with the Declaration of Helsinki.

## Data availability

All data needed to support the conclusions of this study are present in the paper and/or the Supplementary Materials or are available via the corresponding authors upon reasonable request. Values for all data points are available in the Supporting Data Values file.

## ACKOWLEDGEMENTS

This work was supported by the French national research agency (ANR) (Programme d’Investissements d’Avenir ANR-16-IDEX-0007 [NeXT Initiative], ANR-21-CE17-0006 [to RBo], ANR-21-CE14-0016 [to A-CV], and ANR-15-CE17-0008-01 and ANR-23-CE14-0001 [to GL]), Fondation pour la Recherche Médicale (R22131NN - RAD22168NNA to GL), Institut de France – Académie des Sciences (Lamonica Award to GL, supporting M-AM), the Inserm and Regional Council of Pays de la Loire (to RB) and the local fund Genavie (to MF and RR). Study benefited of the framework of the AneuX project supported by grants from SystemsX.ch, and evaluated by the Swiss National Science Foundation (2014/261) exploiting the infrastructure previously elaborated during the @neurIST project supported by the 6th framework program of the European Commission (FP6-IST-2004-027703). We are grateful to the genomics (GenoA), bioinformatics (BiRD) and microscopy core facilities (MicroPICell) (SFR Bonamy, Nantes, France), members of Biogenouest and IBISA, for their expert services. We also thank the *Institut Français de Bioinformatique* (IFB; ANR-11-INBS-0013) as well as the animal house facility (UTE) of Nantes Université. We would like to thank the Genome Aggregation Database (gnomAD) and the groups that provided exome and genome variant data to this resource. A full list of contributing groups can be found at http://gnomad.broadinstitute.org/about. We acknowledge the Center of Biological Resources (CHU Nantes, Hôtel-Dieu, CBR, Nantes, France) as well as Martine Le Cunff and Marie-France Le Cunff and the Clinical Investigation Center 1413 of Nantes for their assistance in managing the ICAN and PREGO biobanks.

## AUTHOR CONTRIBUTIONS

RB, GL and RR designed the research. SCh, EB, SJ, AC, CD, JO, HD, RB and ICAN Study group were involved in the recruitment of IA patients. SM, BRK and PhB, and provided human IA samples. Genetic data have been obtained and analyzed by RB, SBe, SBo, PL, FL, J-FD, CD and RR. MF, M-AM, SPRB, MR, TQ, MB and A-CV performed CTSO function and expression experiments and analyzed the results. SCu performed atomic force microscopy experiments. MF. and RB shared the first-author position for their major contribution to the study: M.F for functional analysis and R.B. for genetic analyses. GL and RR equally contributed to the management of the project, analysis and interpretation of the data and the writing of the manuscript. All authors discussed the results, red, commented and approved the final manuscript.

