## Supplemental materials for "Identification of rare missense variants reducing cathepsin O secretion in families with intracranial aneurysm"

Table S1 to S4

Figures S1 to S2

Appendices

### SUPPLEMENTAL TABLES

**Table S1. Summary of the filtering steps applied to genetic variants detected by whole-exome sequencing in family A.**

| Filtering Step | Remaining Variants | | |
|---|---|---|---|
| **Individuals** | **II:9** | **III:4** | **III:2** |
| **Raw data** | **35070** | **38109** | **37516** |
| *Avg Depth > 15* | *28247* | *28233* | *28279* |
| *Minimum Genotyping Quality > 80* | *21932* | *18943* | *19185* |
| *Filters related to Mapping Quality* # | *16707* | *14185* | *14313* |
| *No Low Complexity Region & Mappability = 1* | *15883* | *13554* | *13681* |
| **Post-QC data** | **15883** | **13554** | **13681** |
| **Functional variants** | **5598** | **5210** | **5183** |
| **MAF < 0.1% (NFE in gnomAD)** | **153** | **112** | **97** |
| **Shared by the 3 relatives (based on WES)** | **8** | | |
| **Shared by all affected (in IBD haplotypes)** | **1 (*CTSO*)** | | |

Abbreviations: MAF, minor allele frequency; NFE: non-Finnish Europeans; GnomAD: Genome Aggregation Database; IBD, identity by descent.

### Retention Criteria: Mapping Quality > 59 & Strand Odd Ratio < 3 & Fisher Strand < 60 & Quality by Depth > 2 & Mapping Quality Rank Sum ∈ [-2.5,2.5].

**Table S2. Rare non-synonymous variants shared by all affected relatives based on WES.**

| Genomic Position (Hg19) | Gene | Nucleotide change | Protein Consequence | Predicted functional impact | | MAF (NFE in Gnomad) |
|---|---|---|---|---|---|---|
| | | | | SnpEFF | CADD score (phred) | |
| 1:46818608:G>A | *NSUN4* | c.661G>A | p.Val221Met | MODERATE | 26.1 | 7.03.10-5 |
| 3:8669403:C>T | *SSUH2* | c.655G>A | p.Ala219Thr | MODERATE | 13.11 | 1.30.10-4 |
| 4:156847208:C>T | *CTSO* | c.946G>A | p.Val316Ile | MODERATE | 25.8 | 1.97.10-5 |
| 8:19680911:A>G | *INTS10* | c.623A>G | p.Asn208Ser | MODERATE | 11.03 | 1.93.10-4 |
| 9:115181197:T>G | *HSDL2* | c.557T>G | p.Phe186Cys | MODERATE | 27.6 | 3.85.10-4 |
| 11:44129522:T>G | *EXT2* | c.359T>G | p.Met120Arg | MODERATE | 24.3 | 9.07.10-4 |
| 16:4861235:C>T | *GLYR1* | c.1523G>A | p.Arg508His | MODERATE | 27.1 | / |
| 21:34625019:A>G | *IFNAR2* | c.593A>G | p.Asp198Gly | MODERATE | 25.2 | 8.42.10-4 |

Abbreviations: MAF, minor allele frequency; NFE, non-finnish european; CADD, combined annotation dependent depletion; GnomAD, genome aggregation database.

**Table S3: Haplotypes (HG19 coordinates) shared based on IBD analysis between individuals II.2, II.9, III.2, III.4 and III.7, with corresponding numbers of included genes.**

| Chromosome | Start | End | # Genes | |
|---|---|---|---|---|
| **2** | 86433240 | 106551158 | 326 | |
| **4** | 155654421 | 165750529 | 72 | (including *CTSO*) |
| **5** | 48534 | 2103907 | 44 | |
| **14** | 69141384 | 76748441 | 135 | |
| **21** | 39014971 | 43459991 | 59 | |

**Table S4. Primer sequences for RT-qPCR experiments**

| Target gene | Forward primer (5'-3' sequence) | Reverse primer (5'-3' sequence) |
|---|---|---|
| *Gapdh* | AACCCATCACCATCTTCCAG | CCAGTAGACTCCACGACATAC |
| *Ctso* | CCAGATACCCAGCAAAAGGA’ | ATCGCACCGCTGACTCTACT |
| *Acta2* | ACGCGAAGCTCGTTATAGAAG | GACCCTGAAGTATCCGATAGAAC |
| *Cnn1* | GATCCACTCTCTCAGCTCCT | CTTCCGCACACTTTAACCGA |
| *Myh11* | CTTTCCAGCTCCAGACTCAC | CGCCTCACATCTATGCCATT |
| *Tagln* | GCTCCTCATCATACTTCTTCTCA | AACGCTACTCTCCTTCCAG |
| *Bmp4* | ATAAAACGACCATCAGCATTCG | GCCTTTCCAGCAAGTTTGTTC’ |
| *Fn1* | CGAGGTGACAGAGACCACAA | CTGGAGTCAAGCCAGACACA |
| *Col1a* | TACAGCACGCTTGTGGATGG | CAGATTGGGATGGAGGGAGTT |

### SUPPLEMENTAL FIGURES

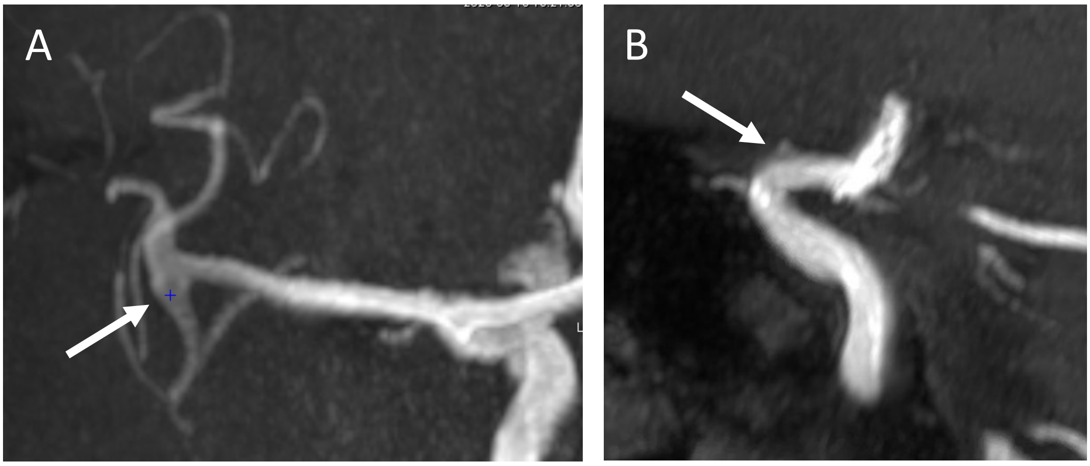


**Figure S1. Magnetic resonance angiographies for two subjects with uncertain phenotypes.** **A,** Fusiform dysplasia located on the middle cerebral artery bifurcation (white arrow) of the subject III-14 in family A. **B,** Ectasia located at the dorsal aspect of the C2 segment on the internal carotid artery (white arrow) of the individual III-9 in family B.

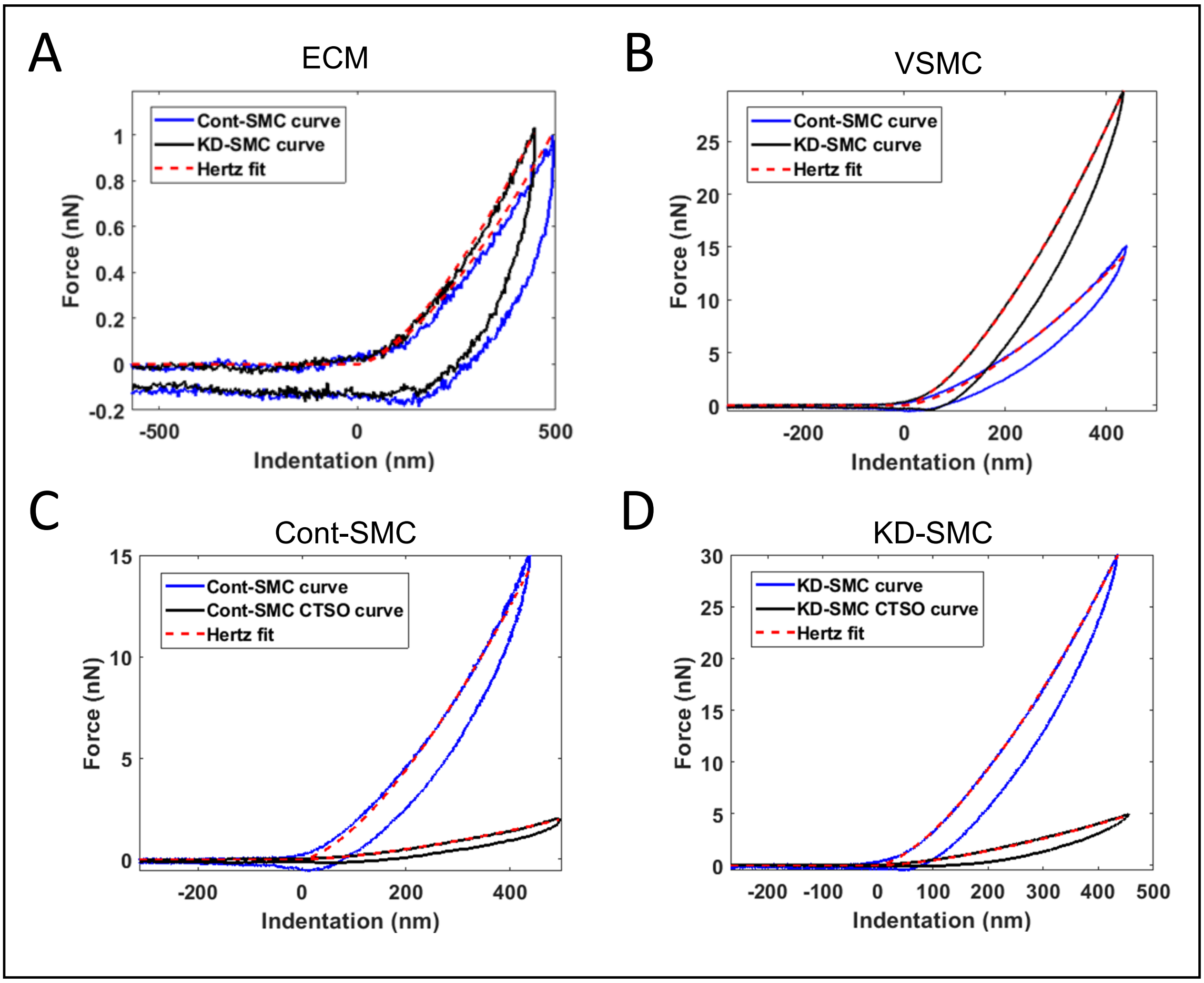


**Figure S2. Analysis of VSMC and ECM by atomic force microscopy.**
**A,** Representative force-indentation curves of decellularized ECM produced by Cont-SMC (blue) and KD-SMC (black). **B,** Representative experimental approach-retract force-indentation curves performed on the cytoplasmic region of Cont-SMC (blue) and KD-SMC (black). **C,** and **D,** Representative experimental approach-retract force-indentation curves performed on the cytoplasmic region of Cont-SMC (**C**) and KD-SMC (**D**) in the absence (blue) and in the presence (black) of CTSO (200 ng/mL added 24 h prior to AFM experiments). The red curves correspond to the Hertz model fit of the approach curve to determine the apparent elastic modulus.

### Appendices

#### Consortia

The ICAN Study Group includes the following investigators: Hubert Desal, Romain Bourcier, Benjamin Daumas-Duport, Bertrand Isidor, Jérôme Connault, Pierre Lebranchu, Thierry Le Tourneau, Marie Pierre Viarouge, Chrisanthi Papagiannaki, Michel Piotin, Hocine Redjem, Mikael Mazighi, Jean Philippe Desilles, Olivier Naggara, Christine Rodriguez, Waghi Ben Hassen, Suzanna Saleme, Charbel Mounayer, Olivier Levrier, Pierre Aguettaz, Xavier Combaz, Anne Pasco, Emeline Berthier, Marc Bintner, Marc Molho, Pascale Gauthier, Cyril Chivot, Vincent Costalat, Cyril Darganzil, Alain Bonafé, Anne Christine Januel, Caterina Michelozzi, Christophe Cognard, Fabrice Bonneville, Philippe Tall, Jean Darcourt, Alessandra Biondi, Cristina Iosif, Elisa Pomero, Jean Chistophe Ferre, Jean Yves Gauvrit, François Eugene, Hélène Raoult, Jean Christophe Gentric, Julien Ognard, René Anxionnat, Serge Bracard, Anne Laure Derelle, Romain Tonnelet, Laurent Spelle, Léon Ikka, Aymeric Rouchaud, Augustin Ozanne, Gildaz Caroff, Nidal Ben Achour, Jacques Moret, Emmanuel Chabert, Jérôme Berge, Guillaume Marnat, Xavier Barreau, Florent Gariel, Frédéric Clarencon, Mohammed Aggour, Frédéric Ricolfi, Adrien Chavent, Pierre Thoant, Brivael Lemogne, Denis Herbreteau, Richard Bibi, Anna Paula Narata, Pedro Lebedenski, Laurent Pierot, Sébastien Soize, Marc Antoine Labeyrie, Christophe Vandendries, Emmanuel Houdart, Appoline Kazemi, Xavier Leclerc, Jean Pierre Pruvo, Sophie Gallas, Stéphane Velasco.
